# Red blood cell distribution width is an indicator of physiological dysregulation and reveals the relationship between biomarkers and health status in hemodialysis patients

**DOI:** 10.64898/2026.09.04.26362306

**Authors:** Yuichi Nakazato, Masahiro Shimoyama, Hirofumi Shimoyama, Hiroaki Kobayashi, Hiroyuki Takao, Hiromi Shimoyama

## Abstract

**Introduction:** Red blood cell distribution width (RDW) and intra-individual variability (IIV) of biomarkers are different types of variation. Although their high levels are commonly associated with high mortality risks and poor clinical outcomes, the reason for this remains unclear.

**Methods:** We explored the significance of this similarity using time-series data (from 143,475 samples) collected over a 9-year period from 1,216 patients on long-term hemodialysis. The IIV of each of the 22 blood-based biomarkers was assessed using the moving coefficient of variation (mCV). Non-linear longitudinal/temporal changes of the variables and their mutual relationships were analyzed using generalized additive (mixed) models.

**Results:** RDW and all of the IIVs showed similar trends throughout the entire period of dialysis treatment: a rapid decrease after hemodialysis initiation followed by an increase that accelerated prior to death. The slopes of these downward and upward trends during each one-year period were statistically different from zero for RDW and the majority of the IIVs. Given the reported increase in the RDW and IIVs of several biomarkers in the pre-dialysis stages of chronic kidney disease, these shared changes might reflect alterations in homeostatic capacity. The RDW exhibited positive correlations with the mCVs of all 22 biomarkers (all p-values < 0.0009), suggesting that it represents dysregulation of the overall physiological system. Furthermore, the levels of the biomarkers showed a specific (nearly linear or U-shaped) relationship with the RDW, analogous to their reported relationship to all-cause mortality in the hemodialysis population.

**Conclusion:** Increased RDW could serve as a marker of physiological dysregulation, and measurement of the RDW can be an effective tool for assessing the overall health status and identifying optimal ranges for various health indicators.

## Introduction

Red blood cell distribution width (RDW) is a measure of the degree of variations in the red blood cell volume (anisocytosis) and was originally used as an adjunct to the diagnosis of anemia. Subsequently, numerous studies have been conducted on various disease cohorts and general populations, revealing that elevated RDW is consistently and strongly associated with an increased risk of death and other adverse outcomes ^1,2^. Furthermore, it has been reported that RDW is also associated with clinical outcomes following therapeutic interventions in diverse medical settings ^3,4^. Although there are several theories regarding why RDW is consistently associated with poor health outcomes across a wide range of clinical situations, the underlying mechanisms remain unclear ^5^.

While the RDW is a variability determined in a single blood test, the variability observed in repeated measurements of physiological/biochemical parameters, referred to as “visit-to-visit variability” or “intra-individual variability” (IIV), has also been reported to be associated with poor health outcomes. At first, increased IIV of the blood pressure or blood glucose was reported to be associated with organ damage, cardiovascular events, and mortality ^6,7^. Subsequently, similar associations have been demonstrated for IIV of numerous blood-based biomarkers and other health parameters, including the blood levels of hemoglobin (Hb), albumin (Alb), creatinine (Cr) and lipids, heart rate, body weight, sleep duration, reaction time, and gait speed ^8,9^.

In patients with end-stage renal disease (ESRD) undergoing maintenance HD therapy, various blood or physiological parameters are measured regularly as part of routine clinical care. Using multivariate high-frequency time-series data recorded during this therapy, we demonstrated that the IIVs of various blood and hemodynamics biomarkers are positively correlated with each other, suggesting that they share information related to physiological dysregulation ^10,11^. Cohen et al. analyzed the data from a cohort of HD patients in Canada and reported that all IIVs for serum sodium (Na), potassium (K), and complete blood count (CBC) parameters increased synchronously prior to death ^12^. In the same study, they demonstrated that the RDW also increased prior to death. Therefore, previous studies suggest that (a) even if the biomarkers are different, their IIVs share common characteristics, and that (b) there are similarities between the IIVs of biomarkers and the RDW levels.

To explore the physiological significance underlining the similarities between these two distinct types of variability, we examined their dynamics and correlations using time-series data that included a broader range of biomarkers.

## Methods

### Study population

A total of 1,337 ESRD patients underwent chronic HD between February 1, 2015 and December 31, 2024 (study period) at any of four affiliated HD facilities in Saitama-City, Japan. Among them, 1,216 patients underwent HD for six months or longer during this period. All of these patients were included as study participants and their demographic and clinical data collected during the study period were used in this study.

Comprehensive written informed consent for use of the clinical data in future research was obtained from all patients at the initial treatment visit to the participating facilities. Furthermore, an opt-out disclosure was provided via posters and on our website in 2024 to ensure that the patients were informed of the study and had the opportunity to refuse participation. This retrospective observational study was conducted in accordance with the Declaration of Helsinki, with the approval of the institutional ethics committee of Hakuyukai Medical Corporation (approval number: 06-002).

### Blood-based and hemodynamic biomarkers

At the four facilities, a total of 26 blood parameters were regularly measured in all patients according to a standardized protocol (Table 1). Blood samples were collected twice monthly, typically at the first HD session of the week (Monday or Tuesday) and were sent to a single external laboratory. In addition, the blood level of glycated albumin (GA) was measured once a month in patients with diabetes mellitus.

**Table 1.**
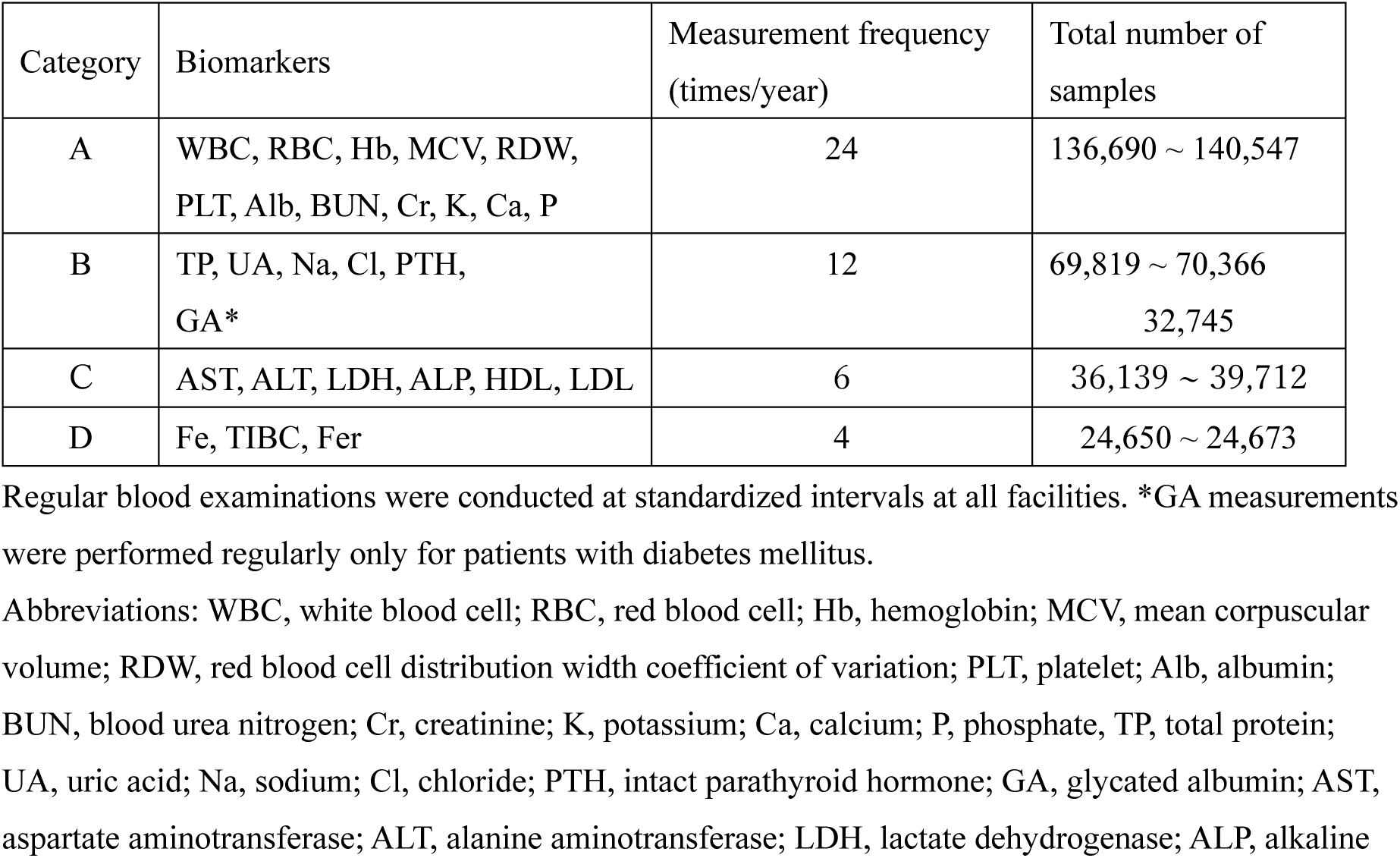
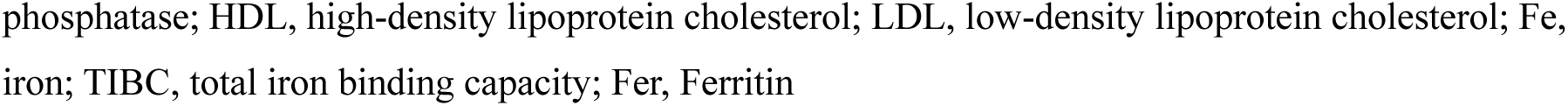
Regular blood examinations.

| Category | Biomarkers | Measurement frequency<br>(times/year) | Total number of<br>samples |
| --- | --- | --- | --- |
| A | WBC, RBC, Hb, MCV, RDW,<br>PLT, Alb, BUN, Cr, K, Ca, P | 24 | 136,690 ~ 140,547 |
| B | TP, UA, Na, Cl, PTH,<br>GA* | 12 | 69,819 ~ 70,366<br>32,745 |
| C | AST, ALT, LDH, ALP, HDL, LDL | 6 | 36,139 ~ 39,712 |
| D | Fe, TIBC, Fer | 4 | 24,650 ~ 24,673 |

Regular blood examinations were conducted at standardized intervals at all facilities. \*GA measurements were performed regularly only for patients with diabetes mellitus.
Abbreviations: WBC, white blood cell; RBC, red blood cell; Hb, hemoglobin; MCV, mean corpuscular volume; RDW, red blood cell distribution width coefficient of variation; PLT, platelet; Alb, albumin; BUN, blood urea nitrogen; Cr, creatinine; K, potassium; Ca, calcium; P, phosphate; TP, total protein; UA, uric acid; Na, sodium; Cl, chloride; PTH, intact parathyroid hormone; GA, glycated albumin; AST, aspartate aminotransferase; ALT, alanine aminotransferase; LDH, lactate dehydrogenase; ALP, alkaline
phosphatase; HDL, high-density lipoprotein cholesterol; LDL, low-density lipoprotein cholesterol; Fe, iron; TIBC, total iron binding capacity; Fer, Ferritin

Data from a small number of ad hoc blood tests were also included in the analysis. The biochemical and hematological data derived from 143,475 blood samples were extracted from the electronic medical records. For ALP measurements, since Japanese medical institutions transitioned from the JSCC to the IFCC method in 2020, previously measured ALP values were converted to IFCC values using a linear conversion formula developed in-house by our medical corporation.

Additionally, we used the pre-dialysis blood pressure and pulse rate data for the year 2020, which had been electronically recorded at two of the four facilities. These data were derived from 343 of the 717 participants in that year and had been analyzed in our previous study ^11^.

### Statistical analysis

All the statistical analyses and data visualizations were performed using R 4.5.1 (R Core Team, 2025) with the gplots, tsModel, psych, car, ggplot2, mgcv, gamm4, itsadug, tidyverse, bestNormalize, and qgraph packages. Results are generally presented as means ± standard deviation (SD), and two-tailed *P* values of < 0.05 were considered as denoting statistical significance.

### Biomarker variability

Intra-individual variability (IIV) of biomarkers was evaluated in two ways.

(1) Coefficient of variation (CV)

IIV in the one-year time-series data (typically 6-24 samples/patient) for each of the 22 blood-based biomarkers (test items for Categories A–C in Table 1, excluding PTH and RDW) were evaluated for each patient using the CV (= population SD/mean). Since the CV for all biomarkers exhibited non-normal distributions with right-skewed tails, the values were log _10_-transformed for the statistical analyses ^13,14^. The log CV value for biomarker X is abbreviated as X-LCV, and the annual mean value of biomarker X is abbreviated as X-M.

(2) Moving CV

In addition to the annual analysis, moving coefficient of variation (mCV) was used to track the biomarker variability over time without restriction to specific time periods. In this study, the mCV was calculated for a moving window containing three consecutive measurements using the formula mCV = moving SD/moving average, and was treated as variability on the middle day ^14^. The small window size (3 measurements) yields unstable mCV values but enables evaluation of the variability at each time point (momentary variability). Even so, when the number of significant digits in the recorded values is small (e.g., Alb, K), the calculated mCV values will be discrete rather than continuous and may even be zero when three consecutive measurements are identical. Such a situation hinders the transformation process for normalizing the mCV (e.g., logarithmic transformation, Box-Cox transformation). To address this issue, we added a small error (20% of the minimum inter-data difference) to the original data using the jitter function and then applied ordered quantile normalization transformation (orderNorm function in bestNormalize package) to the mCV computed from the jittered data. The mean value of the normalized mCV (abbreviated as nmCV) values was 0, with a standard deviation of 1.

### Regression Analysis

The trend of a variable was estimated using an unstandardized slope coefficient (*b* coefficient) of the linear regression model. Slopes calculated for individual patients were subjected to a one-sample t-test to determine if the mean value differed significantly from zero. Non-linear relationships between the variables were modeled using a generalized additive model (GAM) via the gam function in the mgcv package where smoothing parameters are optimized using the default generalized cross validation method. Non-linear temporal changes of the variables (RDW or nmCVs) were estimated using a generalized additive mixed model (GAMM) including the time interval as a fixed effect and subject ID as a random effect. GAMMs were fitted using the gamm4 package, and the parameters were estimated by the restricted maximum likelihood method ^15^.

## Results

### Patient characteristics

Data of a total of 1,216 patients were reviewed in this study. As shown in Table 2, 71.5% of the patients were men, and 47.5% had underlying diabetes mellitus. The patients were classified into the following four groups: (a) a patient group in which ongoing HD treatment was continued throughout the entire study period (n=451); (b) a patient group that had been on HD from prior to the study period but died or dropped out during the study period (n=244); (c) a patient group that was initiated on HD during the study period and then continued thereafter (n=438); and (d) a patient group that was initiated on HD during the study period but dropped out or died during the study period (n=83). Thus, most of the enrolled patients were on long-term HD treatment, and the number of participants in each month during the study period ranged from 604 to 672. From 2015 to 2023, the participants’ average age increased monotonically from 65.7 to 67.7 years, with the overall mean age at the time of all blood samplings being 66.7 ± 12.6 years. Routine blood examinations were conducted according to a common protocol across all facilities (Table 1), and the average number of blood tests each patient underwent during the study period was 118.0 ± 73.3.

**Table 2.** Characteristics of the study population.

|  |  |
| --- | --- |
| Total number of patients, n | 1216 |
| Age* (years), mean ± SD | $66.7 \pm 12.6$ |
| Age* range (years) | 22.1 - 100.5 |
| Men, n (%) | 870 (71.5%) |
| Diabetics, n (%) | 578 (47.5%) |
| HD duration* (years), median (IQR) | 7.7 (3.4 - 14.5) |
| Follow-up length within the study period (years), median (IQR) | 4.2 (2.0 - 8.0) |
| Final HD duration in the study period (years), median (IQR) | 7.9 (3.8 - 15.1) |
| Patients who started HD during the study period, n | 521 |
| Death, n (%) | 327 (26.9%) |
| Age at death (years), median (IQR) | 76.9 (68.9 - 96.1) |
| HD duration at death (years), median (IQR) | 8.8 (4.9 - 16.3) |
| Number of blood tests per patient, median (IQR) | 108 (50 - 196) |

HD, hemodialysis; IQR, interquartile range; \*Calculated by aggregating the patient's age and HD duration at each blood sampling.

### Statistical characteristics of the RDW

Complete Blood Count (CBC), including RDW-CV (abbreviated as RDW), was performed twice monthly, and the monthly average of all the RDW measurements obtained from the study participants is shown in Supplementary Fig. S1-A. Blood samples were initially analyzed by one external laboratory (Lab A), but after October 2016, the analyzing laboratory was switched to a different laboratory (Lab B). While both laboratories used Sysmex hematology analyzers (models unknown) for CBC, the reported RDW values by the two laboratories were significantly different, possibly due to the use of different models. Clear seasonal variations in the RDW were observed, with the RDW peaking in August and reaching its lowest value in January. In addition, an overall upward trend was observed throughout the entire study period, likely reflective of the aging patient population. Based on a linear regression model incorporating the laboratory switch and seasonal variations, 0.45% was subtracted from the RDW values reported by Lab A to calibrate the differences between the two laboratories and the adjusted RDW values were used for the subsequent analyses.

As previously reported ^16^, the RDW exhibited a highly right-skewed distribution and the median value was 13.75% (IQR 13.1-14.6%) (Supplementary Fig. S1-B). The RDW values were Box-Cox transformed to approximate a normal distribution, and the transformed values were denoted as bcRDW. bcRDW = (RDW^λ^ -1)/λ, where λ = -3.7

The mean bcRDW value was 0.2701539 ± 0.0000050. Comparison of the values in men and women of the same age revealed slightly lower average RDW values in women. In a generalized additive model (GAM) with age as a smooth predictor, the sex difference in the predicted bcRDW at age 66.7 (mean age) was equivalent to 0.018% on the RDW scale, indicating that the difference in the RDW between male and female patients was essentially negligible. In the present study, the bcRDW was generally used for correlation analyses and model fitting, but in some graphs, the RDW is intentionally displayed instead of bcRDW to prioritize readability.

### Changes in the RDW after initiation of HD and prior to death in the study patients

Fig. 1 shows the actual RDW data recorded for a patient. Although there is some variation in consecutively measured values, an overall trend can be observed, as indicated by the spline curve fitted to the data. For each patient in whom the HD therapy was started during the study period, the patient’s smoothed spline curve was plotted with aligned time axes (Fig. 2-A). The endpoint of the curve for the patient who died within 5 years of HD initiation is indicated by a black dot. In most cases, the RDW decreased after HD initiation, but began to rise gradually thereafter. Furthermore, in patients who died, the RDW tended to rise more rapidly prior to death. When the trajectories in the deceased patients were plotted with their dates of death aligned, a mostly increasing trend was observed prior to death, regardless of the HD vintage. (Fig. 2-B).

**Fig. 1.**
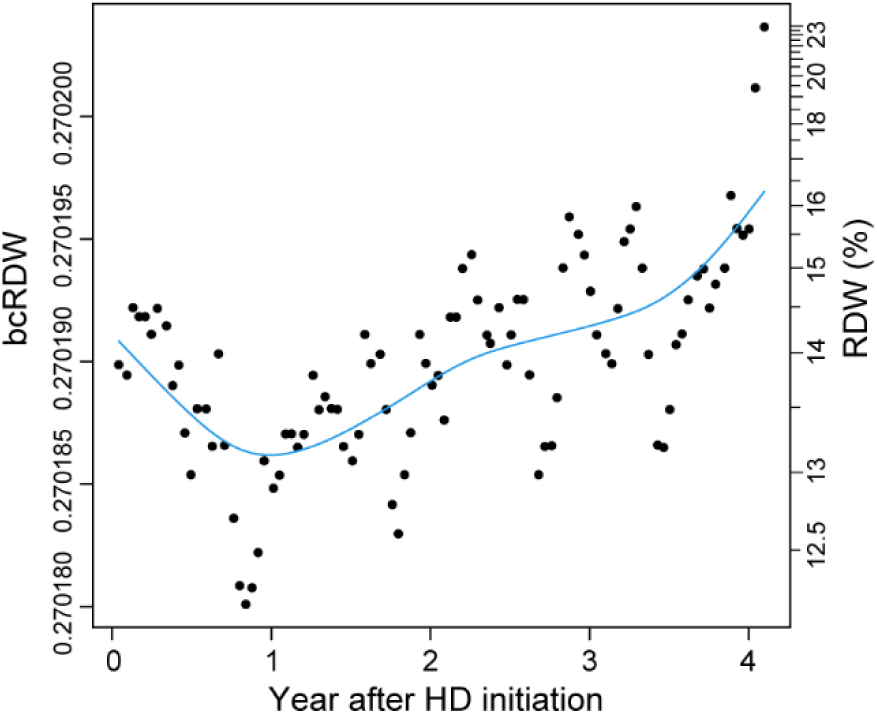
An example of the RDW time series data. Box-Cox transformed RDW (bcRDW) values of a patient were plotted over the 4 years following the initiation of maintenance HD. The blue curve depicts the fitted cubic smoothing spline model. The Y-axis on the right displays the RDW scale corresponding to the bcRDW value.

**Fig. 2.**
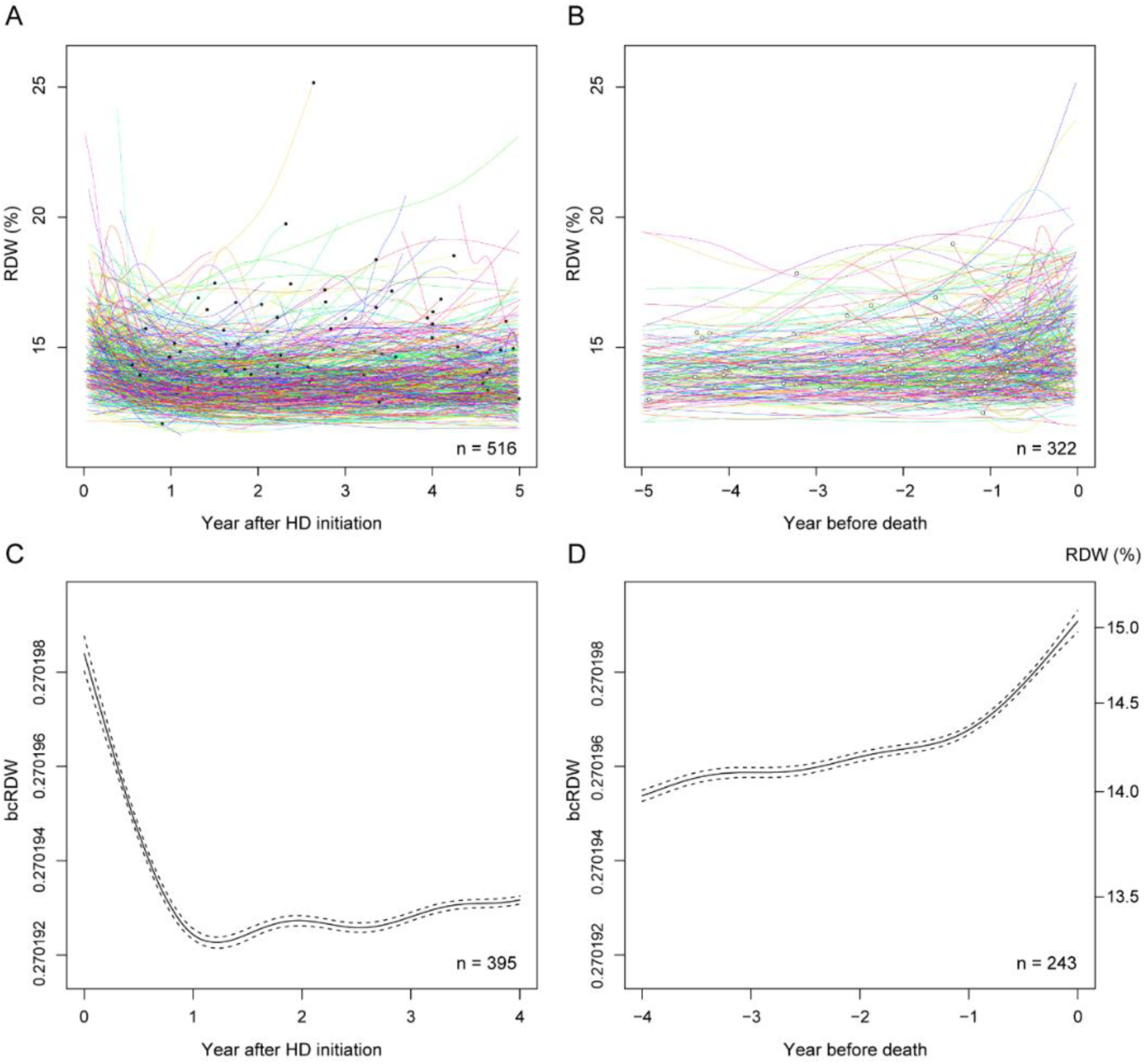
Longitudinal changes of the RDW after the initiation of maintenance HD therapy and before death. (**A**) Individual smoothed trajectories of the RDW values in 516 patients who were initiated on HD therapy during the study period. The filled circles indicate the final predicted RDW values for deceased patients. (**B**) RDW trajectories in 322 patients who died during the study period. The open circles indicate the first HD treatment at any of the participating facilities. (**C**) Overall trend of the RDW following HD initiation was evaluated using a GAMM based on RDW measurements obtained from 395 patients over a 6-year period. (**D**) Similarly, the pre-mortem trend was assessed using RDW measurements obtained from 243 patients over a 6-year period before death. The dashed lines represent the 95% confidence interval.

The overall dynamics of the RDW during the initiation phase of HD and pre-mortem phase were explored using GAMMs (Fig. 2-C, D). The RDW decreased during the first year after HD initiation and began to increase gradually thereafter. The RDW values were generally high in the pre-death period, and with an accelerated increase observed during the last year prior to death.

### Dynamics of biomarker variability after the initiation of HD and before death

We evaluated the IIV for each of 22 biomarkers using its respective normalized moving coefficient of variation (nmCV) and examined its overall dynamics in the same manner as for the RDW (see Methods). As shown in Fig. 3A, the IIV for each biomarker (nmCV) decreased after the initiation of dialysis.

**Fig. 3.**
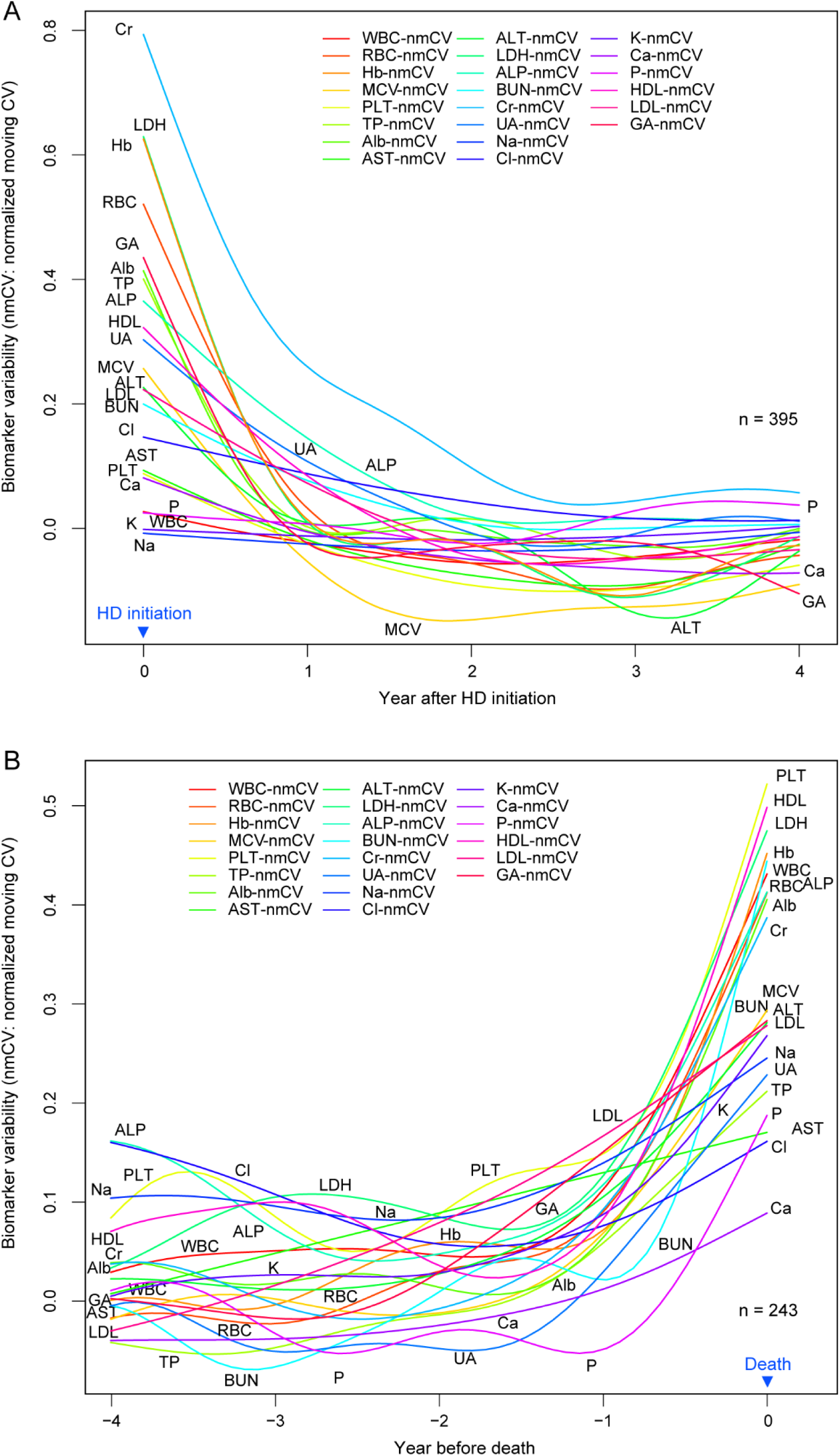
Overall changes in biomarker variabilities after HD initiation and before death. For each of the 22 biomarkers, the momentary variability was estimated by determining the normalized moving CV (nmCV), and the dynamics of the variability was evaluated by fitting GAMM to the nmCV values. In the legends accompanying the curves, the suffix “-nmCV” is omitted. (**A**) Overall trends in biomarker variabilities following HD initiation (N = 395). (**B**) Overall trends in biomarker variabilities before death (N = 243). The patient populations analyzed were the same as those whose data are shown in Fig. 2-C and 2-D.

Subsequently, although the course varied depending on the biomarkers, a trend towards slight increase was observed by 1 to 3 years after the start of dialysis. Furthermore, a synchronized rapid increase was observed in all biomarker variabilities approximately 1 to 2 years prior to death (Fig. 3B). Fig. 2-C, D and Fig. 3-A, B make it clear that the longitudinal changes in the variabilities of both (bcRDW and 22 nmCVs) are similar.

To statistically test these changes observed during the first year after the start of dialysis and during the last year before death, we performed a linear regression analysis on the one-year data from patients who had undergone HD treatment for more than 3 years (Supplementary Table S1). For the first year after HD initiation, the slopes of the regression lines (*b* coefficients) were all negative, and these downward movements were statistically significant for bcRDW and 17 of the 22 nmCVs. For the last year before death, the slopes were all positive and the upward movements were statistically significant for bcRDW and 13 of the 22 nmCVs. In this analysis, the *b* coefficients calculated for frequently measured biomarkers (Category A in Table 1) showed relatively little variation, and most were significantly different from zero, suggesting that the number of data points available for the regression analysis can affect the results.

### Trend of RDW during the maintenance phase of HD therapy

Next, we examined the overall movement of the RDW during the maintenance phase of dialysis, namely, the period between the HD initiation phase and the pre-mortem phase. The total duration of HD varied among patients, and how the maintenance period is defined could influence the results. By referring to Fig. 2, we set the period from one year after HD initiation to one year before death and analyzed the data of 708 patients in whom the maintenance period was two years or longer. Plotting the bcRDW regression lines for each subject according to the age revealed that the bcRDW increased in the majority of cases during this period (Supplementary Fig. S2). The average slope (*b* coefficient) of the individual’s regression lines was 3.83E-7 ± 1.01E-6 and was significantly greater than zero (*P* = 2.22E-16).

### Trend of biomarker variability during the maintenance phase of HD therapy

When a similar analysis was performed on the IIVs for the 22 blood biomarkers (nmCVs), setting the maintenance period became more complex because each biomarker variability appears to reach its minimum at a different time (Fig. 3). Supplementary Table S2 presents the results of a linear regression analysis performed assuming the same maintenance period as in the RDW study: from one year after the start of dialysis until one year before death. For 9 out of the 22 nmCVs, statistically significant upward trends were observed. Thus, not all biomarkers showed an increasing trend in their fluctuations during this period.

### Relationship between RDW and biomarker variability

As RDW and variabilities of a set of biomarkers (22 nmCVs) exhibited largely similar temporal changes throughout the course of HD therapy, we next examined the relationship between the two using a GAM. For all biomarkers except P, the bcRDW showed a nearly linear positive relationship with the biomarker variabilities (Fig. 4). As described in the Methods section, X-nmCV values were calculated from 3 consecutive measurements of biomarker X, which resulted in significant variation between adjacent points. Owing to this variation, the correlation coefficients between the bcRDW and nmCV values were low, although they were all highly significant due to the large number of data points (Supplementary Table S3, right side). The *P* value for P-nmCV was 0.0009, and the values for the nmCVs of all other biomarkers were < 1.1×10^-15^.

**Fig. 4.**
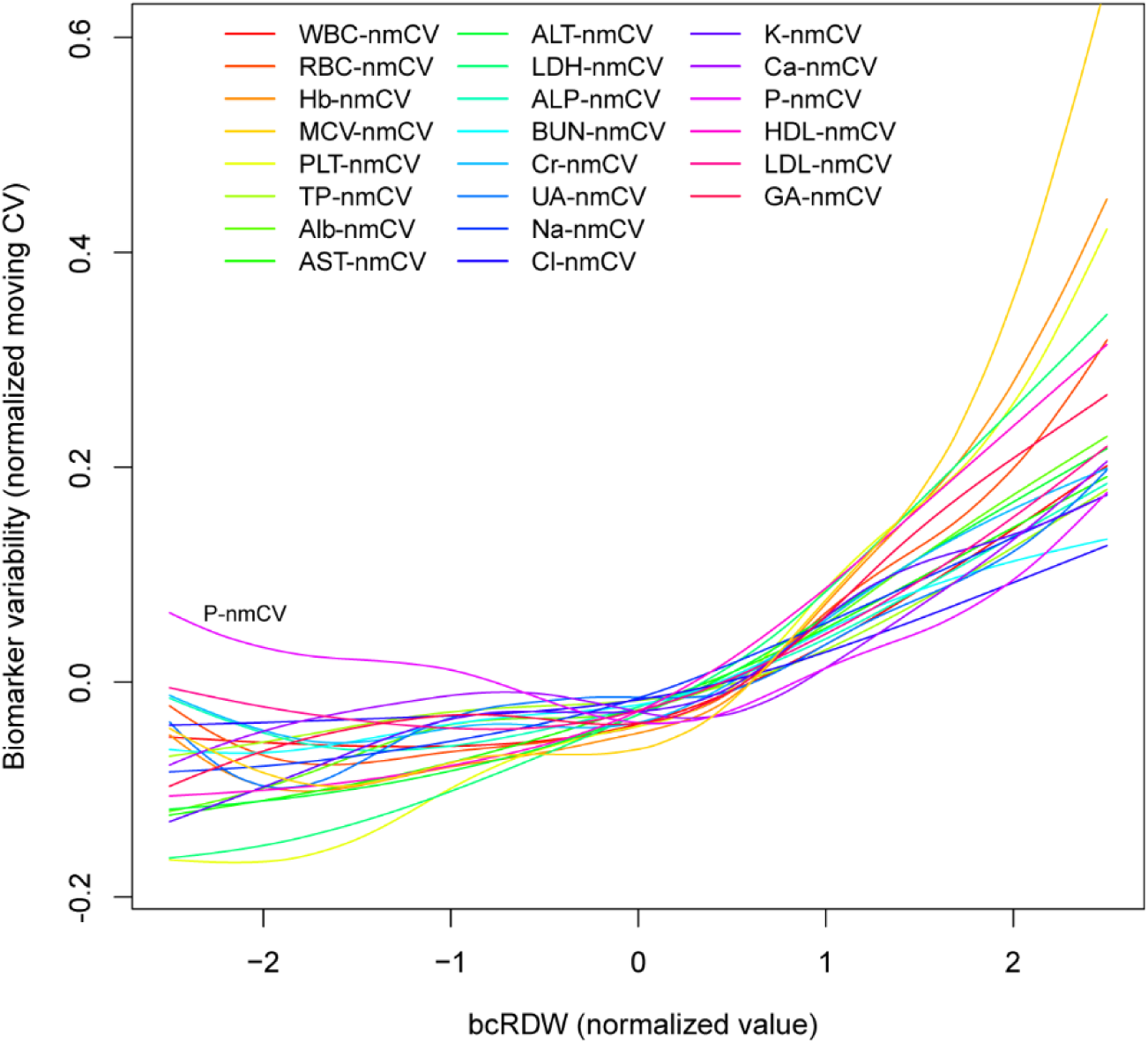
Relationship between the RDW and biomarker variability. Non-linear relationships between the bcRDW and the IIVs of 22 biomarkers (nmCVs) were analyzed using GAMs (N = 29,695 ∼ 134,323).

### RDW in the physiological regulatory system

We have previously reported that there are six regulatory domains within the physiological network formed by the 22 biomarker regulatory systems ^11^. Since the RDW and biomarker IIVs appear to share the same physiological information, we examined whether the RDW might be more closely associated with specific biomarker regulatory systems or domains in the network diagram. To draw the network structure of biomarker regulatory systems, it is necessary to compute a correlation matrix, namely, the correlation coefficients for all pairs of biomarker IIVs. However, not all biomarkers were measured simultaneously (Table 1), which prevented calculation of the correlation matrix. Thus, instead of using nmCV as the IIV estimate for each biomarker, we used log transformed coefficients of variation (LCVs) calculated from the measured data for one year and constructed a correlation matrix for the series of LCVs. Furthermore, since data on the blood pressure (BP) and pulse rate (PR) were only available for the year 2020, the analysis was limited to data from that year. As shown in Table 3 on the right, the annual mean bcRDW (bcRDW-M) showed a positive correlation with the variabilities of all 22 biomarkers (LCVs), with the correlation for 14 of them being statistically significant. The correlation coefficients were relatively uniform, indicating that RDW was associated with all biomarker variabilities in a broad and non-exclusive manner.

**Table 3.** Correlations of the mean bcRDW with biomarker levels/variability based on data from the year 2020.

| Biomarker levels | *r* | *P*-value | Biomarker variabilities | *r* | *P*-value |
| --- | --- | --- | --- | --- | --- |
| ALP-M | 0.194 | **0.0000** | PLT-LCV | 0.296 | **0.0000** |
| AST-M | 0.179 | **0.0000** | Alb-LCV | 0.249 | **0.0000** |
| LDH-M | 0.156 | **0.0001** | Hb-LCV | 0.220 | **0.0000** |
| Cl-M | 0.078 | 0.0588 | WBC-LCV | 0.170 | **0.0000** |
| HDL-M | 0.018 | 0.6553 | DBP-LCV | 0.166 | **0.0020** |
| Na-M | 0.011 | 0.7849 | HDL-LCV | 0.155 | **0.0002** |
| WBC-M | 0.006 | 0.8844 | Na-LCV | 0.154 | **0.0002** |
| ALT-M | -0.010 | 0.8121 | K-LCV | 0.143 | **0.0005** |
| K-M | -0.015 | 0.7169 | LDH-LCV | 0.118 | **0.0042** |
| PLT-M | -0.044 | 0.2891 | BUN-LCV | 0.114 | **0.0055** |
| PR-M | -0.047 | 0.3833 | PR-LCV | 0.114 | **0.0347** |
| Hb-M | -0.050 | 0.2252 | Cr-LCV | 0.090 | **0.0291** |
| TP-M | -0.051 | 0.2129 | Ca-LCV | 0.089 | **0.0310** |
| UA-M | -0.052 | 0.2082 | SBP-LCV | 0.088 | 0.1019 |
| BUN-M | -0.068 | 0.0993 | ALP-LCV | 0.086 | **0.0373** |
| P-M | -0.068 | 0.0976 | UA-LCV | 0.074 | 0.0729 |
| LDL-M | -0.094 | **0.0227** | TP-LCV | 0.073 | 0.0768 |
| SBP-M | -0.101 | 0.0620 | LDL-LCV | 0.071 | 0.0856 |
| Cr-M | -0.190 | **0.0000** | ALT-LCV | 0.060 | 0.1484 |
| DBP-M | -0.235 | **0.0000** | Cl-LCV | 0.058 | 0.1626 |
| Ca-M | -0.269 | **0.0000** | AST-LCV | 0.055 | 0.1853 |
| Alb-M | -0.395 | **0.0000** | P-LCV | 0.011 | 0.7816 |

X-M denotes the mean value of biomarker X calculated from 2020 data, and X-LCV denotes its variability (log-transformed coefficient of variation). The correlation coefficients ( $r$ ) between the mean RDW levels (bcRDW-M) and biomarker levels (Ms) are shown on the left, and the $r$ between the bcRDW-M and biomarker variabilities (LCVs) are shown on the right. The rows in the table are sorted in descending order of the $r$ -values. $P$ values printed in bold indicate $P < 0.05$ . Blood-based data were derived from 589 patients who underwent 21 or more blood chemistry tests during the year, while hemodynamic data (SBP, systolic BP; DBP, diastolic BP; PR) were obtained from 343 patients.

The network diagram (Fig. 5) revealed the presence of six domains (or clusters) in the biomarker regulatory systems as reported previously ^11^, and the RDW did not show any selective linkage with specific biomarker regulatory domain(s). Rather, the RDW was interconnected with the entire system in a balanced manner. The correlations between the bcRDW-M and 22 LCVs, based on one year’s data, were consistent with those obtained from the entire bcRDW and nmCVs dataset (Supplementary Table S3, right side).

**Fig. 5.**
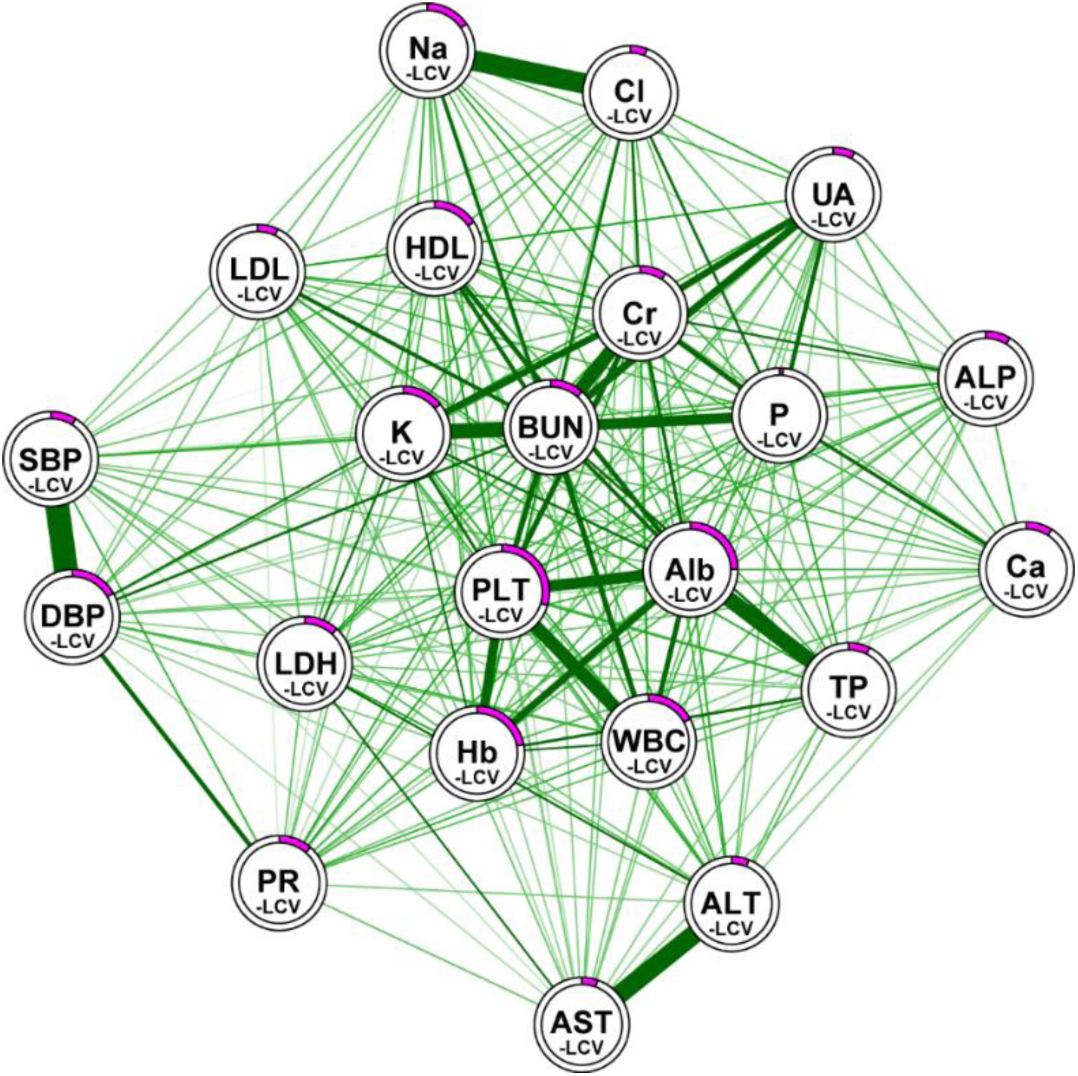
Relationship between the RDW and a biomarker regulatory system. This figure is an extension of the network diagram presented in our previous study (11), incorporating the relationships between RDW and the regulatory mechanisms of each biomarker. Each node labeled X-LCV represents a regulatory system of biomarker X, and the thickness and relative length of the lines connecting the nodes indicate the strength of their correlations. The nodes form six clusters corresponding to the regulatory domains for metabolism, inflammation, circulation, liver, salt, and protein. The magenta band within the ring surrounding each node represents the correlation coefficient between X-LCV and mean bcRDW (bcRDW-M). The correlations were calculated based on data from the year 2020.

### Relationship between the RDW and biomarker levels

These results suggest that an increase in the RDW might serve as a marker of overall dysfunction of the physiological regulatory system, that is, of physiological/homeostatic dysregulation ^10,17,18^. As impaired homeostasis is linked to deteriorating health conditions such as frailty, vulnerability, and fragile resilience ^19^, we examined the relationship between the RDW and biomarker levels. The correlations between the RDW and biomarker levels are shown on the left in Table 3 and Supplementary Table S3. Both tables show negative correlations of the RDW with the serum levels of Alb, Cr, etc., and positive correlations of the RDW with the serum ALP, AST, etc. To examine the non-linear relationships between the RDW and various biomarkers, we further applied these data to univariate GAMs. Figure 6, showing representative results, reveals right-downward relationships of the RDW with the serum Alb, LDL and Cr (panel A-C), and right-upward relationships with the serum AST/ALP and age (panel J-L). In addition, the RDW showed a U-shaped relationship with the Hb, MCV, PLT, Na, transferrin saturation, and GA values (panel D-I).

**Fig. 6.**
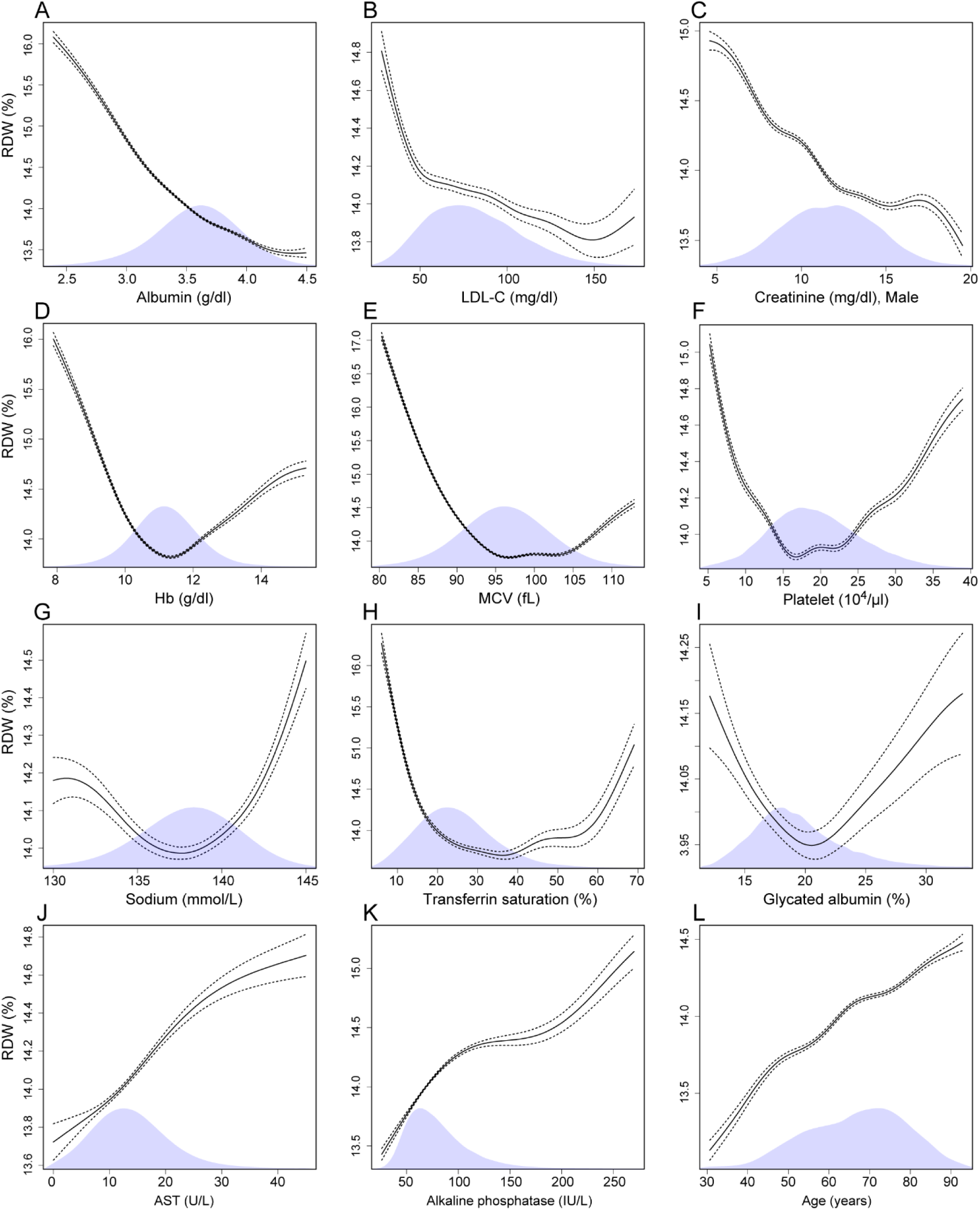
Relationship between biomarkers and the RDW. (A-L) Nonlinear relationships between 12 biomarkers and the RDW were analyzed using univariate GAMs. The results shown are based on data including those from both men and women, except for the serum Cr (C). The dotted lines represent the 95% confidence interval, and the shaded areas denote sample distributions. The number of samples for each model is listed in Supplementary Table S3, on the left side.

Supplementary Fig. S3 summarizes the relationships between available biomarkers and the RDW. For many biomarkers, the regression curve is U-shaped and shows an “optimal range”, indicating that a healthy state is associated with laboratory data being within a specific range.

## Discussion

### Longitudinal changes in the RDW and biomarker variability before death

Among the numerous studies on RDW, only a limited number have examined its changes over time. These studies, focused on patients with sepsis, cardiomyopathy, and heart failure, and ESRD, as well as the elderly, have shown a tendency for the RDW to increase prior to death ^12,20–23^. Based on our results as well, this pattern is likely universal. Similar to the case for the RDW, the IIVs for several biomarkers (Alb, Na, K, and CBC parameters) have also been reported to show an upward trend before death ^12,14^, and in the present study, this upward trend was confirmed across all the 22 biomarkers examined. Our previous research have suggested that increased variability in each biomarker represents deterioration of the respective regulatory system; therefore, the synchronous increase in all biomarker variabilities prior to death indicates that the regulatory dysfunction spread throughout the entire system ^10,11^. In critical transition theory, increased variability indicates system instability and is considered a key early warning signal preceding a critical transition. Our results appear to be in good agreement with this theory ^12,24,25^.

### Increased RDW as a marker of physiological dysregulation

The finding that the RDW and IIVs of numerous biomarkers were positively correlated with each other and exhibited similar dynamics suggests that the RDW, as also the IIVs of biomarkers, represents physiological dysregulation. Previous studies have consistently reported elevated RDW levels in the advanced stages of CKD prior to the initiation of dialysis ^16,26–31^. Similarly, higher IIVs have been demonstrated for several biomarkers in patients with advanced CKD prior to the initiation of hemodialysis, including Hb ^32,33^, HbA1c ^34^, eGFR ^35–38^, serum K ^39^, and BP ^40–42^. In addition to these changes in the pre-dialysis phase, we showed in the present study that both the RDW and biomarker IIVs decreased after the initiation of dialysis therapy.

The kidneys play a major role in maintaining fluid, electrolyte, and acid-base balances through urine production, and contribute to the maintenance of homeostasis. Declining renal function in CKD patients leads to a progressive dysregulation of homeostasis, until life is no longer easy to sustain. HD corrects the imbalances in body composition in place of the kidneys, restoring homeostasis in patients with ESRD. In addition, aging is often accompanied by overall decline in physiological functions, which could lead to frailty, a condition characterized by reduced homeostatic reserve ^17,43^. Several studies reported to date have shown elevated RDW values and increased biomarker variability in frail individuals ^10,44–46^. Therefore, it is evident that the RDW and biomarker IIV dynamics in CKD patients correspond to changes in the individuals’ capacity to maintain homeostasis. (refer to the schema in Fig. 7)

**Fig. 7.**
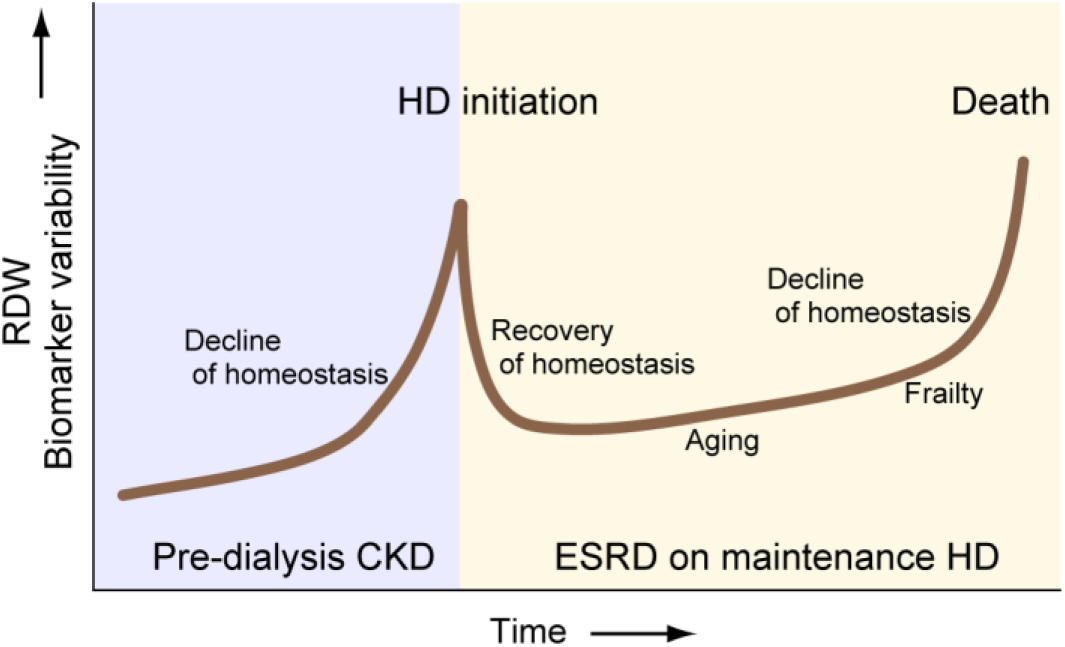
Schematic diagram illustrating the synchronous changes in the RDW and biomarker variability in CKD patients.

### Broad relationship between the RDW and biomarker regulatory systems

Increase in RDW and IIVs of the examined biomarkers can be considered as representing dysregulation, and the strength of their interrelationship is reflected in the correlation coefficients. The series of correlation coefficients between the RDW and biomarker IIVs (Table 3 and Supplementary Table S3, right side) were relatively uniform, with the exception of that for P variability. It has been shown that the P variability (P-LCV) is the highest among LCVs for commonly measured biomarkers ^10,13,47^. This is likely caused by external factors such as changes in diet and ongoing adjustments of medications to control P and PTH levels ^48,49^. In other words, circumstances where the intrinsic P regulatory capacity does not readily translate into P-LCV may underlie the low correlation observed between bcRDW-M and P-LCV.

Homeostasis is thought to be maintained by the cooperation of various regulatory mechanisms within the body ^11^. In a network diagram (Fig. 5), RDW did not show any selective relationship with a specific regulatory domain. Such diffuse associations of the RDW with multiple biomarker regulatory mechanisms indicate the potential utility of RDW as an indicator of system-wide physiological dysregulation ^50^.

### Biomarker-RDW relationships vs. Biomarker-Mortality relationships

RDW is a single metric and whether it can truly evaluate overall physiological dysregulation without bias requires further confirmation. Nonetheless, our conclusion was strongly supported by the results of comparing the known relationships between biomarker levels and all-cause mortality with the biomarker-RDW relationships. Several previous survival analyses in HD patients have revealed negative correlations between the serum Alb, LDL and Cr levels and mortality ^51–53^. As shown on the left in Table 3 and Supplementary Table S3, the levels of these biomarkers also showed negative correlations with the RDW. Serum AST and ALP levels and age, which are known to be positively correlated with the mortality risk ^54–56^ were also positively correlated with the RDW levels. In addition to these monotonic relationships, many other biomarkers are known to exhibit U-shaped relationships with the mortality risk. For example, for biomarkers such as Hb ^57^, MCV ^58^, PLT ^59^, Na ^60,61^, TSAT ^62^, and GA ^63^, specific levels, referred to as being within the “optimal range” are associated with lower mortality risks^64,65^.

Significantly, these biomarker levels also showed quite analogous U-shaped relationships with the RDW (Fig. 6). This close association between the RDW and mortality risk lends itself to the following interpretations: (a) increased RDW indicates physiological dysregulation and ill health; (b) mortality risk is primarily determined by the extent of the physiological dysregulation; (c) changes in biomarker levels may reflect impaired physiological regulation. Homeostatic capacity determines vulnerability, resilience, and frailty, and is essential for sustaining life ^19,66,67^. Therefore, if we consider increase in the RDW as a marker of physiological dysregulation, its associations with ill health, frailty, and poor clinical outcomes can be reasonably explained.

The association between biomarker levels and prognosis varies depending on the population studied; such a phenomenon is known as “reverse epidemiology” or “cholesterol paradox” ^68^. In dialysis patients, different from the general population, lower levels of cholesterol, Cr, BUN, BP, and BMI are associated with worse outcomes ^69,70^. In line with this finding, the RDW was found to show negative correlations with the levels of LDL, Cr, BUN, and BP (Supplementary Table S3 and Table 3). Multiple studies have also reported the existence of negative associations between the serum LDL levels and mortality risk in patients with heart failure ^68,71,72^. Interestingly, Dang et al. reported that the RDW was positively associated with the BUN and Cr levels and negatively associated with the LDL level in heart failure patients ^73^. Thus, the RDW may accurately reflect the relationship between biomarker levels and the health status across different cohorts.

### Applications of RDW in clinical research

Mortality and morbidity rates have long been used to investigate the impact of various health-related factors/indicators (i.e., predictors) on health status. In such studies, known as survival analysis, it is necessary to collect baseline data and then track the occurrence of events over a certain period of time. As stated above, we demonstrated the high similarity between the RDW level and the mortality risk in terms of the relationships with numerous biomarkers. This result suggests that health status can be easily assessed by measuring RDW, without the need for an observation period. This new approach can be applied to any health parameters, as long as it is measured concurrently with RDW in a large population. In addition to the blood biomarkers, other measures—such as bio-impedance readings, ECG parameters, sleep duration—may also be subject to this approach.

There may be some differences between the results obtained from these two approaches. In survival analyses, the occurrence of events is observed over a specific period of time, so that the results might reflect the cumulative effects of past physical conditions. On the other hand, the biomarker-RDW relationships observed in this study presumably reflect contemporaneous biomarker-health relationships. In this regard, we may be able to assess the temporal effect of the physical condition on the subsequent health status by examining the relationships between past biomarker data and the subsequently measured RDW values. In addition to its application in observational studies, RDW may also be useful in intervention studies. By monitoring the RDW values after the start of medical or physical therapy, we may be able to objectively assess its impact on the patients’ overall health and select the optimal therapeutic approach.

### Limitations

There were several limitations of this study that need to be borne in mind. First, although our conclusion regarding the significance of the RDW in our study patients is consistent with the findings observed in other cohorts, our analysis was based solely on data from HD patients. Therefore, further verification in other populations would be desirable to ensure generalizability of our findings. Second, no clear theoretical explanation has been established as to how a single metric like the RDW can come to predict overall physiological dysregulation. Accordingly, at this stage, RDW should be regarded as one of the metrics for evaluating physiological dysregulation, and its characteristics should be further explored by comparing it with other health-related indices such as biological age measures, allostatic load, Mahalanobis distance, frailty index/phenotype, etc. Third, RDW test results may vary depending on the analyzer used. RDW data obtained from different medical institutions cannot be considered as being readily comparable. Fourth, as seen in Fig. 1, the RDW itself can exhibit significant variability in repeated measurements. Therefore, for its use in clinical practice, statistical procedures such as averaging and model fitting based on multiple measurements should be considered. Fifth, the RDW has also been shown to exhibit clear seasonal variation. Therefore, when analyzing changes over a short period of time, it is necessary to take this factor into account.

## Data Availability

The analyses in this study required sets of personally identifiable information, such as the date of birth, date of HD initiation, date of death, and dates of all blood examinations, and it is not possible to de-identify the data while preserving these pieces of information. Therefore, to protect the confidentiality of the subjects and comply with the terms of the patient's consent, the raw data cannot be made openly available. However, the aggregate data supporting our findings in this study are included within the manuscript and Supplementary materials. If additional anonymized aggregated data is required, it will be available upon reasonable request by contacting the corresponding author.

## Author Contributions

Y.N., M.S., Hf.S., H.K., and H.S. designed research; Y.N., Hf.S., and H.T. analyzed data; Y.N. and M.S. wrote the paper; M.S., Hf.S., H.K., H.T., and H.S. collected and organized clinical data.

## References

1. Salvagno GL, Sanchis-Gomar F, Picanza A, Lippi G. Red blood cell distribution width: A simple parameter with multiple clinical applications. Crit Rev Clin Lab Sci. 2015;52(2):86–105. doi:10.3109/10408363.2014.992064

2. Pan J, Borné Y, Engström G. The relationship between red cell distribution width and all-cause and cause-specific mortality in a general population. Sci Rep. 2019;9(1):16208-. doi:10.1038/s41598-019-52708-2

3. Garg R. Beyond anemia: Red cell distribution width as a universal biomarker in contemporary medicine. J Hematol Allied Sci. 2025;5(2):115–124. doi:10.25259/jhas_12_2025

4. Li J, Xu Y, Tan S De, Wang Z. Impact of red blood cell distribution width (RDW) on postoperative outcomes in hepatocellular carcinoma (HCC) patients. Med (United States). 2024;103(24):E38475. doi:10.1097/MD.0000000000038475

5. Li N, Zhou H, Tang Q. Red Blood Cell Distribution Width: A Novel Predictive Indicator for Cardiovascular and Cerebrovascular Diseases. Dis Markers. 2017;2017(1):7089493. 10.1155/2017/7089493

6. Parati G, Stergiou GS, Dolan E, Bilo G. Blood pressure variability: clinical relevance and application. In: Journal of Clinical Hypertension. Vol 20. Blackwell Publishing Inc.; 2018:1133–1137. doi:10.1111/jch.13304

7. Ma WY, Li HY, Pei D, et al. Variability in hemoglobin A1c predicts all-cause mortality in patients with type 2 diabetes. J Diabetes Complications. 2012;26(4):296–300. doi:10.1016/j.jdiacomp.2012.03.028

8. Zhu Y, Lu JM, Yu Z Bin, et al. Intra-individual variability of total cholesterol is associated with cardiovascular disease mortality: A cohort study. Nutr Metab Cardiovasc Dis. 2019;29(11):1205–1213. doi:10.1016/j.numecd.2019.07.007

9. Batterham PJ, Bunce D, Mackinnon AJ, Christensen H. Intra-individual reaction time variability and all-cause mortality over 17 years: A community-based cohort study. Age Ageing. 2014;43(1):84–90. doi:10.1093/ageing/aft116

10. Nakazato Y, Sugiyama T, Ohno R, et al. Estimation of homeostatic dysregulation and frailty using biomarker variability: a principal component analysis of hemodialysis patients. Sci Rep. 2020;10(1):10314. doi:10.1038/s41598-020-66861-6

11. Nakazato Y, Shimoyama M, Cohen AA, et al. Intercorrelated variability in blood and hemodynamic biomarkers reveals physiological network in hemodialysis patients. Sci Rep. 2023;13(1):1660. doi:10.1038/s41598-023-28345-1

12. Cohen AA, Leung DL, Legault V, et al. Synchrony of biomarker variability indicates a critical transition: Application to mortality prediction in hemodialysis. iScience. 2022;25(6):104385. doi:10.1016/j.isci.2022.104385

13. Nakazato Y, Kurane R, Hirose S, Watanabe A, Shimoyama H. Variability of laboratory parameters is associated with frailty markers and predicts non-cardiac mortality in hemodialysis patients. Clin Exp Nephrol. 2015;19(6):1165–1178. doi:10.1007/s10157-015-1108-0

14. Nakazato Y, Kurane R, Hirose S, Watanabe A, Shimoyama H. Aging and death-associated changes in serum albumin variability over the course of chronic hemodialysis treatment. Barretti P, ed. PLoS One. 2017;12(9):e0185216. doi:10.1371/journal.pone.0185216

15. Wood S, Scheipl F. Generalized Additive Mixed Models using “mgcv” and “lme4” [R package gamm4 version 0.2-7]. CRAN Contrib Packag. Published online April 22, 2025. doi:10.32614/CRAN.package.gamm4

16. Yoo KD, Oh HJ, Park S, et al. Red blood cell distribution width as a predictor of mortality among patients regularly visiting the nephrology outpatient clinic. Sci Rep. 2021;11(1):24310-. doi:10.1038/s41598-021-03530-2

17. Li Q, Wang S, Milot E, et al. Homeostatic dysregulation proceeds in parallel in multiple physiological systems. Aging Cell. 2015;14(6):1103–1112. doi:10.1111/acel.12402

18. Arbeev KG, Ukraintseva S V., Bagley O, et al. “Physiological Dysregulation” as a Promising Measure of Robustness and Resilience in Studies of Aging and a New Indicator of Preclinical Disease. Journals Gerontol Ser A. 2019;74(4):462–468. doi:10.1093/gerona/gly136

19. Varadhan R, Seplaki CL, Xue QL, Bandeen-Roche K, Fried LP. Stimulus-response paradigm for characterizing the loss of resilience in homeostatic regulation associated with frailty. Mech Ageing Dev. 2008;129(11):666–670. doi:10.1016/j.mad.2008.09.013

20. He R, Liao Y, Men L, et al. Longitudinal modeling of red blood cell distribution width dynamics and mortality risk in critically Ill patients with sepsis-associated acute kidney injury. PLoS One. 2025;20(10 October):e0333605. doi:10.1371/journal.pone.0333605

21. Núñez J, Núñez E, Rizopoulos D, et al. Red blood cell distribution width is longitudinally associated with mortality and anemia in heart failure patients. Circ J. 2014;78(2):410–418. doi:10.1253/circj.CJ-13-0630

22. Chen S, Nie R, Wang Y, et al. Integrating Dynamic Red Blood Cell Distribution Width Monitoring and β-Blocker Therapy for Mortality Prediction in Intensive Care Unit Cardiomyopathy Patients: A Bayesian Multivariate Joint Model and Machine Learning Study. Diagnostics. 2025;15(10):1236. doi:10.3390/diagnostics15101236

23. Martínez-Velilla N, Cambra-Contin K, García-Baztán A, Alonso-Renedo J, Herce PA, Ibáñez-Beroiz B. Change in red blood cell distribution width during the last years of life in geriatric patients. J Nutr Heal Aging. 2015;19(5):590–594. doi:10.1007/s12603-015-0470-7

24. Scheffer M, Carpenter SR, Lenton TM, et al. Anticipating critical transitions. Science (80- ). 2012;338(6105):344–348. doi:10.1126/science.1225244

25. Dakos V, Bascompte J. Critical slowing down as early warning for the onset of collapse in mutualistic communities. Proc Natl Acad Sci U S A. 2014;111(49):17546–17551. doi:10.1073/pnas.1406326111

26. Lippi G, Targher G, Montagnana M, Salvagno GL, Zoppini G, Guidi GC. Relationship between red blood cell distribution width and kidney function tests in a large cohort of unselected outpatients. Scand J Clin Lab Invest. 2008;68(8):745–748. doi:10.1080/00365510802213550

27. Solak Y, Gaipov A, Turk S, et al. Red Cell Distribution Width Is Independently Related to Endothelial Dysfunction in Patients With Chronic Kidney Disease. Am J Med Sci. 2014;347(2):118–124. doi:10.1097/MAJ.0b013e3182996a96

28. Gu L, Xue S. The association between red blood cell distribution width and the severity of diabetic chronic kidney disease. Int J Gen Med. 2021;14:8355–8363. doi:10.2147/IJGM.S332848

29. Deng X, Gao B, Wang F, Zhao MH, Wang J, Zhang L. Red Blood Cell Distribution Width Is Associated With Adverse Kidney Outcomes in Patients With Chronic Kidney Disease. Front Med. 2022;9:877220. doi:10.3389/fmed.2022.877220

30. Roumeliotis S, Stamou A, Roumeliotis A, et al. Red blood cell distribution width is associated with deterioration of renal function and cardiovascular morbidity and mortality in patients with diabetic kidney disease. Life. 2020;10(11):1–16. doi:10.3390/life10110301

31. Yonemoto S, Hamano T, Fujii N, et al. Red cell distribution width and renal outcome in patients with non-dialysis-dependent chronic kidney disease. PLoS One. 2018;13(6):e0198825. doi:10.1371/journal.pone.0198825

32. Sumida K, Diskin CD, Molnar MZ, et al. Pre-End-Stage Renal Disease Hemoglobin Variability Predicts Post-End-Stage Renal Disease Mortality in Patients Transitioning to Dialysis. Am J Nephrol. 2017;46(5):397–407. doi:10.1159/000484356

33. Szeto CC, Kwan BCH, Chow KM, Pang WF, Leung CB, Li PKT. Haemoglobin variability in Chinese pre-dialysis CKD patients not receiving erythropoietin. Nephrol Dial Transplant. 2011;26(9):2919–2924. doi:10.1093/ndt/gfq824

34. Gupta S, Priya N. Glycemic Variability in Different Stages of Chronic Kidney Disease with Type 2 Diabetes Mellitus: A Cross-sectional Study. Indian J Med Biochem. 2024;28(1):1–7. doi:10.5005/jp-journals-10054-0228

35. Lee S, Park S, Kim Y, et al. Impact of variability in estimated glomerular filtration rate on major clinical outcomes: A nationwide population-based study. PLoS One. 2020;15(12):e0244156. doi:10.1371/journal.pone.0244156

36. Suzuki A, Obi Y, Hayashi T, et al. Visit-to-visit variability in estimated glomerular filtration rate predicts hospitalization and death due to cardiovascular events. Clin Exp Nephrol. 2019;23(5):661–668. doi:10.1007/s10157-019-01695-9

37. Tseng CL, Lafrance JP, Lu SE, et al. Variability in estimated glomerular filtration rate values is a risk factor in chronic kidney disease progression among patients with diabetes. BMC Nephrol. 2015;16(1):34-. doi:10.1186/s12882-015-0025-5

38. Nishiwaki H, Missikpode C, Ricardo AC, et al. Time-Updated Estimated GFR Variability Is Associated With Mortality, Cardiovascular Disease, and End-Stage Kidney Disease in Patients With CKD: Findings From the CRIC Study. Am J Kidney Dis. 2025;85(6):695–703.e1. doi:10.1053/j.ajkd.2025.01.010

39. Hsieh MF, Wu; I-Wen, Lee CC, Wang SY, Wu MS. Higher Serum Potassium Level Associated with Late Stage Chronic Kidney Disease. Chang Gung Med J. 2011;34:418–443.

40. Sarafidis PA, Ruilope LM, Loutradis C, et al. Blood pressure variability increases with advancing chronic kidney disease stage: A cross-sectional analysis of 16 546 hypertensive patients. J Hypertens. 2018;36(5):1076–1085. doi:10.1097/HJH.0000000000001670

41. Andersson U, Nilsson PM, Kjellgren K, et al. Variability in home blood pressure and its association with renal function and pulse pressure in patients with treated hypertension in primary care. J Hum Hypertens. 2024;38(3):212–220. doi:10.1038/s41371-023-00874-2

42. Jeffers BW, Zhou D. Relationship between Visit-to-Visit Blood Pressure Variability (BPV) and Kidney Function in Patients with Hypertension. Kidney Blood Press Res. 2017;42(4):697–707. doi:10.1159/000484103

43. Fried LP, Cohen AA, Xue QL, Walston J, Bandeen-Roche K, Varadhan R. The physical frailty syndrome as a transition from homeostatic symphony to cacophony. Nat Aging. 2021;1(1):36–46. doi:10.1038/s43587-020-00017-z

44. Li CM, Chao C Ter, Chen SI, Han DS, Huang KC. Elevated Red Cell Distribution Width Is Independently Associated With a Higher Frailty Risk Among 2,932 Community-Dwelling Older Adults. Front Med. 2020;7:1–7. doi:10.3389/fmed.2020.00470

45. Kim KM, Lui LY, Browner WS, et al. Association Between Variation in Red Cell Size and Multiple Aging-Related Outcomes. J Gerontol A Biol Sci Med Sci. 2021;76(7):1288–1294. doi:10.1093/gerona/glaa217

46. Fravel MA, Ernst ME, Woods RL, et al. Long-term blood pressure variability and frailty risk in older adults. J Hypertens. 2024;42(2):244–251. doi:10.1097/HJH.0000000000003599

47. Levitt H, Smith KG, Rosner MH. Variability in calcium, phosphorus, and parathyroid hormone in patients on hemodialysis. Hemodial Int. 2009;13(4):518–525. doi:10.1111/j.1542-4758.2009.00393.x

48. Leung S, McCormick B, Wagner J, et al. Meal phosphate variability does not support fixed dose phosphate binder schedules for patients treated with peritoneal dialysis: A prospective cohort study. BMC Nephrol. 2015;16(1):205. doi:10.1186/s12882-015-0205-3

49. Tao X, Zhang H, Yang Y, Zhang C, Wang M. Daily dietary phosphorus intake variability and hemodialysis patient adherence to phosphate binder therapy. Hemodial Int. 2019;23(4):458–465. doi:10.1111/hdi.12769

50. Gross AL, Carlson MC, Chu NM, et al. Derivation of a measure of physiological multisystem dysregulation: Results from WHAS and health ABC. Mech Ageing Dev. 2020;188:111258. doi:10.1016/j.mad.2020.111258

51. Sanz-García C, Rodríguez-García M, Górriz-Teruel JL, et al. Differences in association between hypoalbuminaemia and mortality among younger versus older patients on haemodialysis. Clin Kidney J. 2025;18(1). doi:10.1093/CKJ/SFAE339

52. Song JH, Park EH, Bae J, et al. Effect of low-density lipoprotein level and mortality in older incident statin-naïve hemodialysis patients. BMC Nephrol. 2023;24(1):289-. doi:10.1186/s12882-023-03337-5

53. Sakao Y, Ojima T, Yasuda H, et al. Serum creatinine modifies associations between body mass index and mortality and morbidity in prevalent hemodialysis patients. PLoS One. 2016;11(3):e0150003. doi:10.1371/journal.pone.0150003

54. Ling S, Diao H, Lu G, Shi L. Associations between serum levels of liver function biomarkers and all-cause and cause-specific mortality: a prospective cohort study. BMC Public Health. 2024;24(1):3302-. doi:10.1186/s12889-024-20773-6

55. Chang JF, Feng YF, Peng YS, et al. Combined Alkaline Phosphatase and Phosphorus Levels as a Predictor of Mortality in Maintenance Hemodialysis Patients. Medicine (Baltimore). 2014;93(18):e106. doi:10.1097/MD.0000000000000106

56. Blayney MJ, Pisoni RL, Bragg-Gresham JL, et al. High alkaline phosphatase levels in hemodialysis patients are associated with higher risk of hospitalization and death. Kidney Int. 2008;74(5):655–663. doi:10.1038/ki.2008.248

57. Kosugi T, Hasegawa T, Imaizumi T, et al. Association between hemoglobin level and mortality in patients undergoing maintenance hemodialysis: a nationwide dialysis registry in Japan. Clin Exp Nephrol. 2025;29(6):831–842. doi:10.1007/s10157-025-02632-9

58. Honda H, Kimachi M, Kurita N, Joki N, Nangaku M. Low rather than high mean corpuscular volume is associated with mortality in Japanese patients under hemodialysis. Sci Rep. 2020;10(1):15663-. doi:10.1038/s41598-020-72765-2

59. Zhao X, Karaboyas A, Gan L, et al. Platelet count has a U-shaped association with mortality in hemodialysis patients. Sci Rep. 2024;14(1):26572. doi:10.1038/s41598-024-77718-7

60. Chen S, Pan B, Lou X, Chen J, Zhang P. Effect of long-term serum sodium levels on the prognosis of patients on maintenance hemodialysis. Ren Fail. 2024;46(1):2314629. doi:10.1080/0886022X.2024.2314629

61. Rhee CM, Ravel VA, Ayus JC, et al. Pre-dialysis serum sodium and mortality in a national incident hemodialysis cohort. Nephrol Dial Transplant. 2016;31(6):992–1001. doi:10.1093/ndt/gfv341

62. Kuo KL, Liu JS, Lin MH, et al. Association of anemia and iron parameters with mortality among prevalent peritoneal dialysis patients in Taiwan: the AIM-PD study. Sci Rep. 2022;12(1):1269-. doi:10.1038/s41598-022-05200-3

63. Hoshino J, Abe M, Hamano T, et al. Glycated albumin and hemoglobin A1c levels and cause-specific mortality by patients’ conditions among hemodialysis patients with diabetes: A 3-year nationwide cohort study. BMJ Open Diabetes Res Care. 2020;8(1):1642. doi:10.1136/bmjdrc-2020-001642

64. Wen S, Zhou S, Wang W, Qiu X, Feng Y. Associations Between Hematologic Parameters and All-Cause Death in Individuals With Cardio-Renal-Metabolic Multimorbidity: A National Cohort Study. J Am Heart Assoc. 2025;14(18):e041978. doi:10.1161/JAHA.125.041978

65. Halma M, Aniwar M, Selem E, Tuszynski J, Varon J, Marik P. Blood biomarkers associated with all-cause mortality risk: Accessibility and clinical utility for lifestyle medicine. Arch Gerontol Geriatr Plus. 2025;2(2):100145. doi:10.1016/j.aggp.2025.100145

66. Ramsay DS, Woods SC. Clarifying the roles of homeostasis and allostasis in physiological regulation. Psychol Rev. 2014;121(2):225–247. doi:10.1037/a0035942

67. Kotas ME, Medzhitov R. Homeostasis, Inflammation, and Disease Susceptibility. Cell. 2015;160(5):816–827. doi:10.1016/j.cell.2015.02.010

68. Kalantar-Zadeh K, Block G, Horwich T, Fonarow GC. Reverse epidemiology of conventional cardiovascular risk factors in patients with chronic heart failure. J Am Coll Cardiol. 2004;43(8):1439–1444. doi:10.1016/j.jacc.2003.11.039

69. Kalantar-Zadeh K, Ikizler TA, Block G, Avram MM, Kopple JD. Malnutrition-Inflammation Complex Syndrome in Dialysis Patients: Causes and Consequences. Am J Kidney Dis. 2003;42(5):864–881. doi:10.1016/j.ajkd.2003.07.016

70. Georgianos PI, Agarwal R. Blood pressure and mortality in long-term hemodialysis-time to move forward. Am J Hypertens. 2017;30(3):211–222. doi:10.1093/ajh/hpw114

71. Wang Q, Liu Z, Zhou W, et al. LDL cholesterol and clinical outcomes in heart failure with reduced ejection fraction a competing risk analysis. Sci Rep. 2025;15(1):40939-. doi:10.1038/s41598-025-24832-9

72. Gouveia R, Madureira S, Elias C, et al. Lower low density lipoprotein cholesterol associates to higher mortality in non-diabetic heart failure patients. Int J Cardiol Cardiovasc Risk Prev. 2023;18:200197. doi:10.1016/j.ijcrp.2023.200197

73. Dang HNN, Viet Luong T, Cao MTT, Bui VT, Tran TT, Nguyen HM. Assessing red blood cell distribution width in Vietnamese heart failure patients: A cross-sectional study. Klisic A, ed. PLoS One. 2024;19(7):e0301319. doi:10.1371/journal.pone.0301319

