## Supplemental Materials for "Red blood cell distribution width is an indicator of physiological dysregulation and reveals the relationship between biomarkers and health status in hemodialysis patients"

Author affiliations:
[a]Division of Nephrology, Yuai Clinic, Hakuyukai Medical Corporation, Saitama-City, Saitama 330-0854, Japan
[b]Division of Nephrology, Yuai Nisshin Clinic, Hakuyukai Medical Corporation, Saitama-City, Saitama 331-0823, Japan
[c]Division of Nephrology, Yuai Mihashi Clinic, Hakuyukai Medical Corporation, Saitama-City, Saitama 330-0856, Japan
[d]Division of Nephrology, Yuai Minuma Clinic, Hakuyukai Medical Corporation, Saitama-City, Saitama 337-0041, Japan

**Corresponding author**:
Yuichi Nakazato


**This PDF file includes:**

**Supplementary Figures S1 to S3**
**Supplementary Tables S1 to S3**

**Figures**

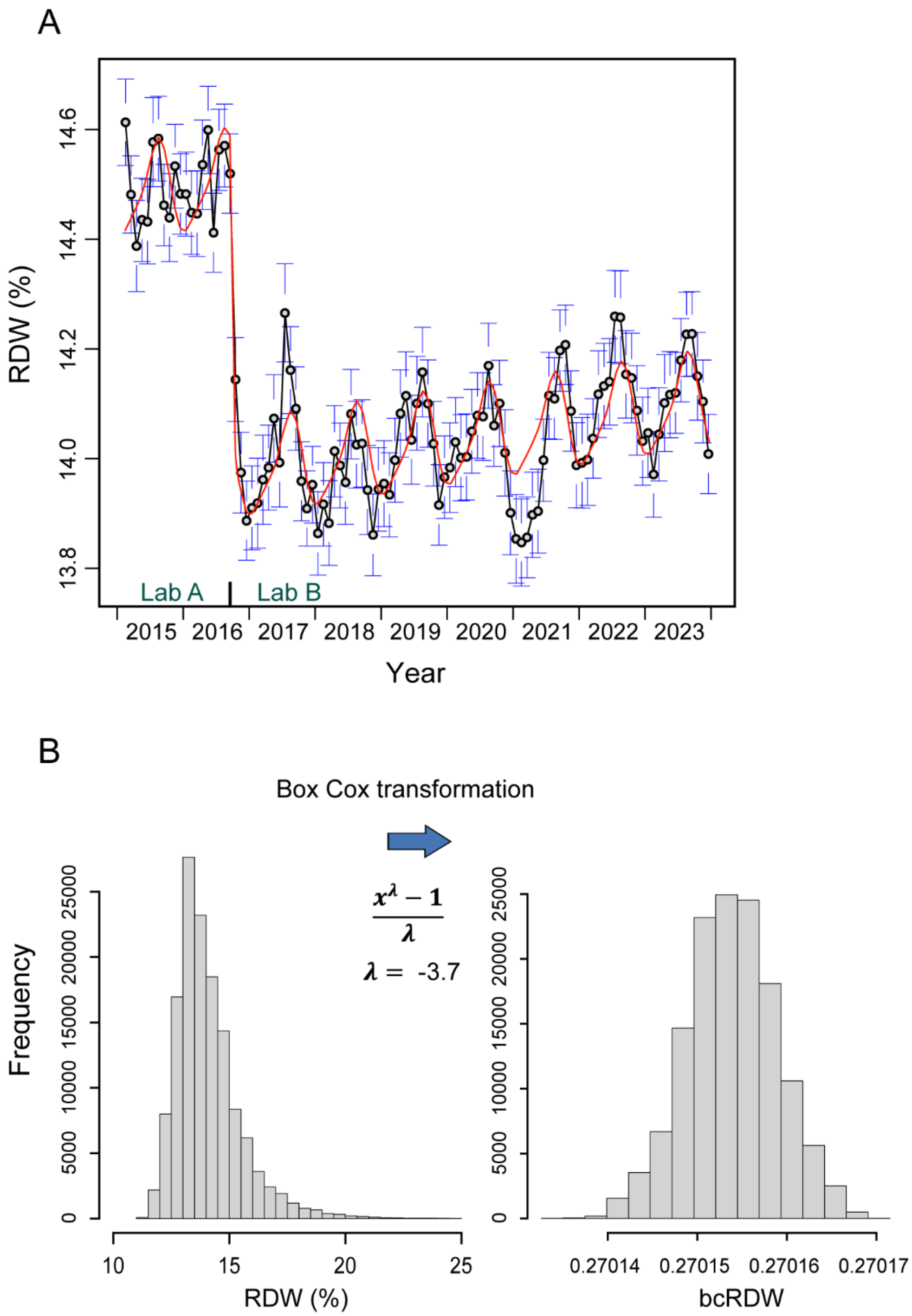


**Fig. S1. Changes in the monthly average value of the RDW during the study period and the distribution of the RDW measurements.** (A) All RDW measurements obtained from the study participants were averaged for each month and plotted with bars representing 95% confidence intervals. The red curve represents the fitted linear regression model incorporating the laboratory switch and seasonal variations. (B) Distribution of the RDW values adjusted for the inter-laboratory difference and that of the Box-Cox transformed RDW (bcRDW) values (N = 136,690).

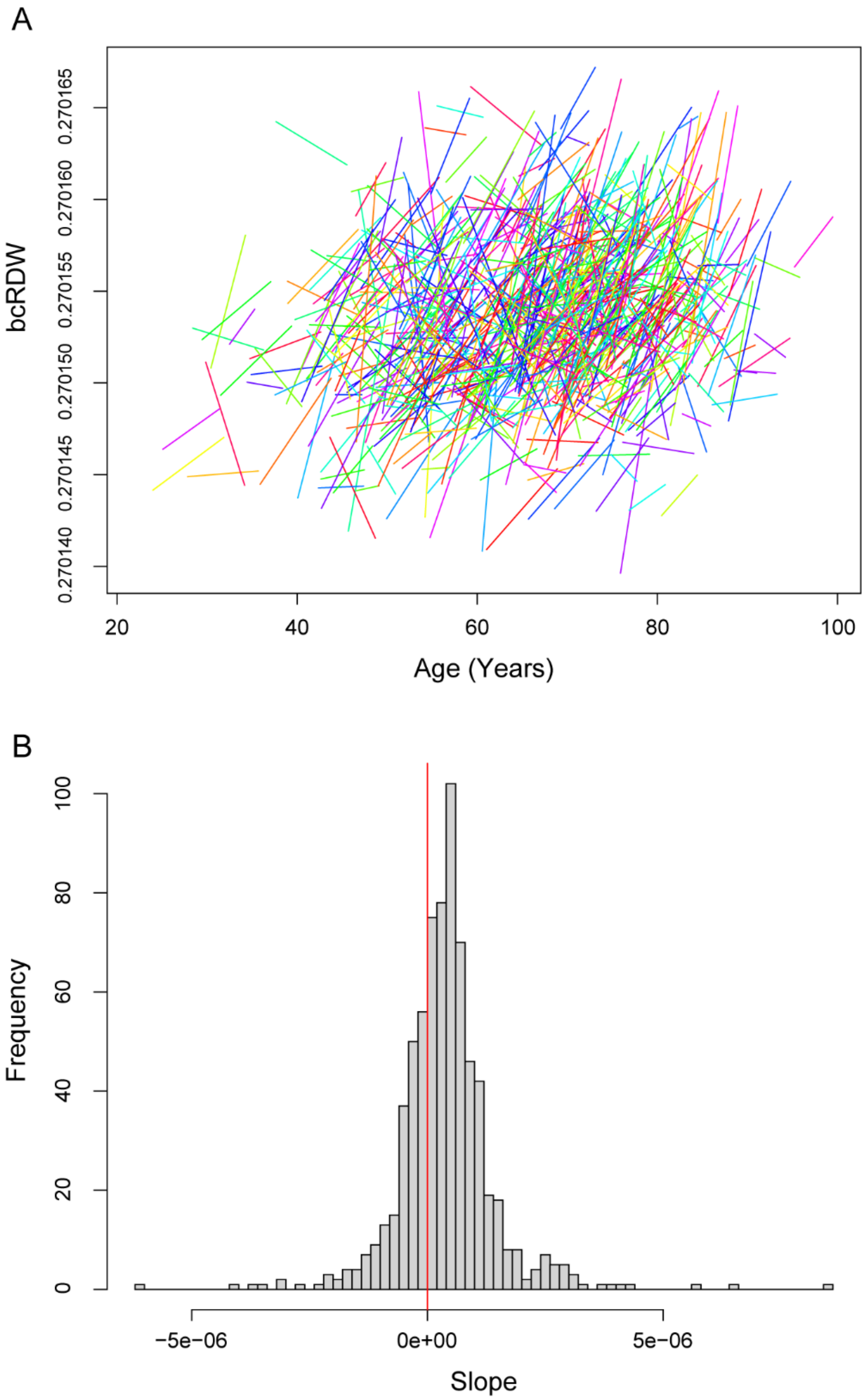


**Fig. S2. Trends in individual RDW values during the maintenance phase of HD therapy.** (A) For each eligible patient, a regression line was calculated based on the RDW values measured during the maintenance period and was placed according to the age. (B) Distribution of the slopes of the regression lines (N = 708).

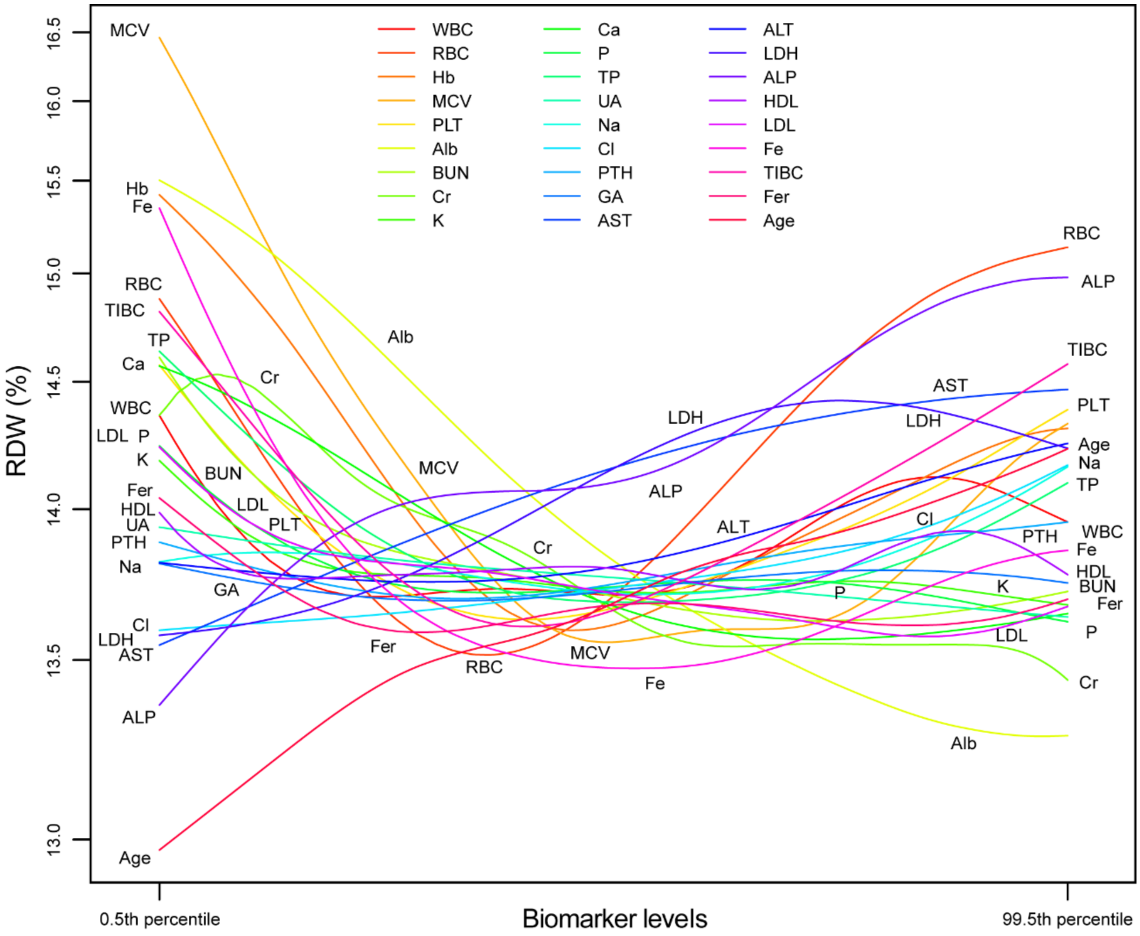


**Fig. S3. Summary diagram demonstrating the relationships between the RDW and 27 biomarkers.** The impact of each biomarker level on the RDW was visualized using univariate GAM and plotted within the range of the 0.5th percentile to the 99.5th percentile of all measured values.

### Tables

**Table S1. Slopes of the regression lines for biomarker variability during the first year of HD therapy and during the last year before death**

| First year after HD initiation (N=143) | | | | | Last year before death (N=201) | | | | |
|---|---|---|---|---|---|---|---|---|---|
| Variable | Slope | | | *P*-value | Variable | Slope | | | *P*-value |
| bcRDW* | -1.24 | ± | 1.22 | **0.000** | bcRDW* | 0.57 | ± | 0.99 | **0.000** |
| WBC-nmCV | -0.15 | ± | 1.10 | 0.095 | WBC-nmCV | 0.35 | ± | 1.26 | **0.000** |
| RBC-nmCV | -0.52 | ± | 1.11 | **0.000** | RBC-nmCV | 0.31 | ± | 1.20 | **0.000** |
| Hb-nmCV | -0.55 | ± | 1.11 | **0.000** | Hb-nmCV | 0.37 | ± | 1.17 | **0.000** |
| MCV-nmCV | -0.38 | ± | 1.01 | **0.000** | MCV-nmCV | 0.18 | ± | 1.15 | **0.032** |
| PLT-nmCV | -0.30 | ± | 1.06 | **0.001** | PLT-nmCV | 0.44 | ± | 1.19 | **0.000** |
| TP-nmCV | -0.42 | ± | 1.47 | **0.001** | TP-nmCV | 0.07 | ± | 1.73 | 0.560 |
| Alb-nmCV | -0.54 | ± | 1.09 | **0.000** | Alb-nmCV | 0.42 | ± | 1.19 | **0.000** |
| AST-nmCV | -0.14 | ± | 1.75 | 0.329 | AST-nmCV | 0.01 | ± | 2.18 | 0.970 |
| ALT-nmCV | -0.22 | ± | 1.70 | 0.119 | ALT-nmCV | 0.14 | ± | 2.22 | 0.390 |
| LDH-nmCV | -0.81 | ± | 1.89 | **0.000** | LDH-nmCV | 0.18 | ± | 2.21 | 0.246 |
| ALP-nmCV | -0.36 | ± | 1.75 | **0.015** | ALP-nmCV | 0.18 | ± | 2.24 | 0.267 |
| BUN-nmCV | -0.41 | ± | 1.10 | **0.000** | BUN-nmCV | 0.41 | ± | 1.28 | **0.000** |
| Cr-nmCV | -0.59 | ± | 1.08 | **0.000** | Cr-nmCV | 0.36 | ± | 1.19 | **0.000** |
| UA-nmCV | -0.62 | ± | 1.64 | **0.000** | UA-nmCV | 0.17 | ± | 1.72 | 0.154 |
| Na-nmCV | -0.30 | ± | 1.56 | **0.024** | Na-nmCV | 0.07 | ± | 1.45 | 0.478 |
| Cl-nmCV | -0.31 | ± | 1.48 | **0.012** | Cl-nmCV | 0.15 | ± | 1.49 | 0.150 |
| K-nmCV | -0.17 | ± | 1.12 | 0.076 | K-nmCV | 0.19 | ± | 1.26 | **0.036** |
| Ca-nmCV | -0.15 | ± | 1.24 | 0.140 | Ca-nmCV | 0.17 | ± | 1.12 | **0.033** |
| P-nmCV | -0.18 | ± | 1.00 | **0.037** | P-nmCV | 0.24 | ± | 1.33 | **0.012** |
| HDL-nmCV | -1.00 | ± | 1.99 | **0.000** | HDL-nmCV | 0.58 | ± | 2.53 | **0.002** |
| LDL-nmCV | -0.58 | ± | 1.95 | **0.001** | LDL-nmCV | 0.31 | ± | 1.99 | **0.032** |
| GA-nmCV | -0.54 | ± | 1.50 | **0.000** | GA-nmCV | 0.05 | ± | 1.60 | 0.760 |

For each variable (bcRDW or nmCV), and for each patient, a linear regression model was built based on measured data for one year. The *b* coefficients of the models are given as slopes. Eligible patients were those who underwent 21 or more blood tests during the first year after HD initiation or during the year prior to death, and those who had undergone continuous HD therapy for at least three years. The analysis for the first year of HD therapy was limited to patients who enrolled within 45 days after HD initiation. *Standardized bcRDW values (Z-scores) were used for the linear regression. *P* values printed in bold indicate $P < 0.05$.

**Table S2. Trends in the RDW and biomarker variabilities during the maintenance phase of HD therapy**

| Variables | Slope (mean ± SD) | *P*-value |
|---|---|---|
| bcRDW* | 0.077 ± 0.219 | **0.000** |
| WBC-nmCV | 0.012 ± 0.173 | 0.068 |
| RBC-nmCV | 0.015 ± 0.165 | **0.014** |
| Hb-nmCV | 0.014 ± 0.158 | **0.018** |
| MCV-nmCV | 0.030 ± 0.147 | **0.000** |
| PLT-nmCV | 0.024 ± 0.187 | **0.001** |
| TP-nmCV | 0.011 ± 0.233 | 0.205 |
| Alb-nmCV | 0.022 ± 0.166 | **0.001** |
| AST-nmCV | 0.028 ± 0.257 | **0.003** |
| ALT-nmCV | 0.016 ± 0.316 | 0.173 |
| LDH-nmCV | 0.005 ± 0.259 | 0.600 |
| ALP-nmCV | -0.021 ± 0.262 | **0.031** |
| BUN-nmCV | 0.010 ± 0.154 | 0.076 |
| Cr-nmCV | -0.010 ± 0.174 | 0.127 |
| UA-nmCV | -0.003 ± 0.206 | 0.657 |
| Na-nmCV | 0.009 ± 0.211 | 0.245 |
| Cl-nmCV | -0.013 ± 0.210 | 0.090 |
| K-nmCV | 0.018 ± 0.166 | **0.004** |
| Ca-nmCV | 0.022 ± 0.180 | **0.002** |
| P-nmCV | 0.000 ± 0.170 | 0.983 |
| HDL-nmCV | 0.005 ± 0.289 | 0.638 |
| LDL-nmCV | 0.021 ± 0.273 | **0.037** |
| GA-nmCV | 0.034 ± 0.403 | 0.131 |

*Standardized bcRDW values (Z-scores) were used for the linear regression. For each variable (bcRDW or nmCV), and for each patient, a linear regression model was built from data within the maintenance period of HD therapy (from one year after the start of dialysis until one year before death). The regression coefficient *b* of the model is referred to as the slope (N = 708). *P* values printed in bold indicate $P < 0.05$.

**Table S3. Correlations of the bcRDW with biomarker levels/variability**

| Biomarker levels | N* | *r* | *P*-value | Biomarker variabilities | N* | *r* | *P*-value |
|---|---|---|---|---|---|---|---|
| ALP | 38,401 | 0.155 | **0.0000** | PLT-nmCV | 13,4323 | 0.100 | **0.0000** |
| RBC | 136,688 | 0.148 | **0.0000** | LDH-nmCV | 35,303 | 0.098 | **0.0000** |
| LDH | 37,842 | 0.127 | **0.0000** | MCV-nmCV | 134,323 | 0.096 | **0.0000** |
| Cl | 69,473 | 0.079 | **0.0000** | Hb-nmCV | 134,323 | 0.085 | **0.0000** |
| AST | 38,624 | 0.078 | **0.0000** | HDL-nmCV | 332,59 | 0.084 | **0.0000** |
| Na | 69,473 | 0.029 | **0.0000** | ALT-nmCV | 359,62 | 0.071 | **0.0000** |
| WBC | 136,687 | 0.019 | **0.0000** | RBC-nmCV | 1343,23 | 0.065 | **0.0000** |
| ALT | 38,526 | 0.013 | **0.0113** | AST-nmCV | 36,096 | 0.063 | **0.0000** |
| PLT | 136,688 | 0.000 | 0.9482 | Alb-nmCV | 134,257 | 0.058 | **0.0000** |
| HDL | 35,903 | -0.004 | 0.5032 | GA-nmCV | 29,695 | 0.057 | **0.0000** |
| GA | 32,387 | -0.011 | **0.0482** | Na-nmCV | 67,027 | 0.054 | **0.0000** |
| K | 136,683 | -0.036 | **0.0000** | K-nmCV | 134,256 | 0.053 | **0.0000** |
| P | 136,683 | -0.036 | **0.0000** | Cr-nmCV | 134,255 | 0.050 | **0.0000** |
| UA | 69,264 | -0.044 | **0.0000** | WBC-nmCV | 134,322 | 0.049 | **0.0000** |
| TP | 69,647 | -0.055 | **0.0000** | ALP-nmCV | 35,882 | 0.048 | **0.0000** |
| LDL | 35,907 | -0.079 | **0.0000** | LDL-nmCV | 33,260 | 0.044 | **0.0000** |
| Hb | 136,688 | -0.086 | **0.0000** | UA-nmCV | 66,806 | 0.043 | **0.0000** |
| BUN | 136,683 | -0.100 | **0.0000** | BUN-nmCV | 134,255 | 0.043 | **0.0000** |
| Ca | 136,683 | -0.158 | **0.0000** | TP-nmCV | 67,194 | 0.038 | **0.0000** |
| Cr | 136,688 | -0.184 | **0.0000** | Cl-nmCV | 67,027 | 0.032 | **0.0000** |
| MCV | 136,683 | -0.184 | **0.0000** | Ca-nmCV | 134,258 | 0.030 | **0.0000** |
| Alb | 136,683 | -0.306 | **0.0000** | P-nmCV | 134,256 | 0.009 | **0.0009** |

The correlation coefficients (*r*) between the bcRDW and biomarker levels are shown on the left, and those for biomarker variabilities (nmCVs) are shown on the right. The rows in the table are sorted in descending order by the *r* values. *P* values printed in bold indicate $P < 0.05$. *N represents the number of paired samples.
